# Risk of COVID-19 and long-term exposure to air pollution: evidence from the first wave in China

**DOI:** 10.1101/2020.04.21.20073700

**Authors:** Pai Zheng, Yonghong Liu, Hongbin Song, Chieh-Hsi Wu, Bingying Li, Moritz U.G. Kraemer, Huaiyu Tian, Xing Yan, Yuxin Zheng, Nils Chr. Stenseth, Christopher Dye, Guang Jia

**Author notes:** Corresponding author. (G.J.). These authors contributed equally to this work (Pai Zheng, Yonghong Liu, Hongbin Song, Chieh-Hsi Wu).

## Abstract

People with chronic obstructive pulmonary disease, cardiovascular disease or hypertension have a high risk of severe coronavirus disease 2019 (COVID-19). Long-term exposure to air pollution, especially PM_2.5_, has also been associated with COVID-19 mortality. We collated individual-level data of confirmed COVID-19 cases during the first wave of the epidemic in mainland China. We fitted a generalized linear model using city-level COVID-19 cases and severe cases as the outcome, and long-term average levels of air pollutants as the exposure. Our analysis was adjusted using several variables, including a mobile phone dataset, covering human movement from Wuhan before the travel ban and movements within each city during the time of emergency response. Other variables included census, smoking prevalence, climate, and socio-economic data from 324 cities in China. We adjusted for human mobility and socio-economic factors, and found that an increase in long-term NO_2_ or PM_2.5_ may correspond to an increase in the number of COVID-19 cases and severe infections. However, the linkage might also be affected by the confounding factor of population size because of the predefined correlation between population size and air pollution. The results are derived from a large, newly compiled and geocoded repository of population and epidemiological data relevant to COVID-19. The findings of this paper (and other previous studies that have given ambiguous results) indicate that a more definitive analysis is needed of the link between COVID-19 and air pollution.

## Introduction

The novel coronavirus disease (COVID-19) has rapidly spread across the world ^1-4^ As more countries have passed the (first) peak of the COVID-19 epidemic, country-based strategies have shifted from containment to delay and mitigation, aiming to prevent a second epidemic curve or lower its peak ^5^. The virus (SARS-CoV-2) induces respiratory stress, so individuals with a compromised respiratory system are expected to be more vulnerable to infection ^6^, and people with pre-existing conditions are more vulnerable to severe infection. Meta-analysis has shown that chronic obstructive pulmonary disease (COPD), cardiovascular disease (CVD), and hypertension are associated with severe COVID-19 infection and admission to intensive care units (ICUs) ^7^.

Long-term exposure to air pollution affects lung function and is associated with increased prevalence of COPD, cardiovascular disease and hypertension ^8-10^. However, it is not known whether vulnerability to lung diseases can affect COVID-19 risk ^11^. We need to understand the impact of air pollution exposure on COVID-19 infection and disease severity. This could help to improve future modelling and disease burden calculations around the world.

During the first wave of COVID-19 in China, to prevent further dissemination of the illness, Wuhan prohibited all transport in and out of the city on 23 January 2020. In the following days, cities across mainland China activated the highest level of emergency response and contained outbreaks outside Wuhan up until 6 March 2020. These measures substantially reduced air pollution: a significant reduction in pollution level was observed across cities in China between January and March 2020 ^12,13^. Data on cities (excluding Wuhan) are ideal to assess the relationship between long-term average exposure and COVID-19 risk (Figure 1) because the cities had widely distributed COVID-19 cases, highly variable historical air quality, and have been surveyed comprehensively under consistent criteria and data standards across the country. Air pollution has both acute and long-term health effects, so air pollution reduction serves as a natural experiment to offset the potential acute effects in particular.

**Figure 1.**
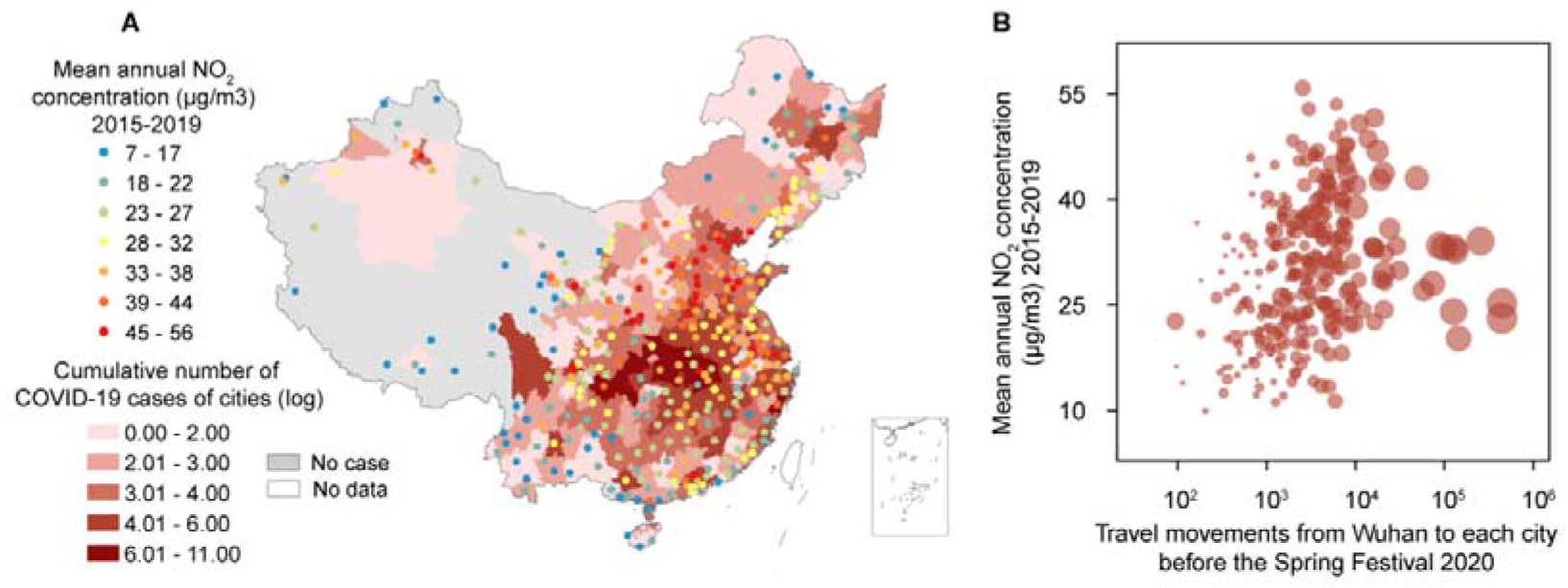
Air pollution exposure, COVID-19, and travel movements in 324 cities of China during Spring Festival 2020. (A) Distribution of cities with data on NO_2_ and COVID-19. The shading from light red to dark red represents the cumulative number of confirmed COVID-19 cases in each city from low to high, from 31 December 2019 to 6 March 2020. Points coloured from blue to red represent the historic mean annual NO_2_ concentration (μg/m^3^) from low to high, during January 2015 and December 2019, before the COVID epidemic. (B) Association between the cumulative number of confirmed cases, the number of human movements from Wuhan to each city, and historic mean annual NO_2_ concentration. The area of circles represents the cumulative number of cases reported by 6 March 2020.

## Methods

### Epidemiological, demographic and geographical data

We collected epidemiological data from the official reports of the health commissions of 324 cities, excluding Wuhan. These included daily reports from 31 December 2019 to 6 March 2020, but excluded newly-reported locally-acquired infections. Data on the percentage of severe COVID-19 cases were obtained from official reports of Provincial Health Committees. The age structure for each city was obtained from the Sixth National Population Census of the People’s Republic of China. Socio-economic data, including the gross domestic product (GDP) per capita in 2016, were obtained from China City Statistical Yearbook.

### Human mobility data

Human movements were tracked using mobile phone data from Baidu location-based services (LBS) and telecommunications operators. The number of recorded movements from Wuhan city to other cities across China was calculated from 11 to 23 January 2020. On 23 January, movement from Wuhan reached almost zero because of the travel ban. Travel movements within each city were recorded until 6 March.

### Data for air pollution

Original daily data for particulate matter concentrations, including PM_2.5_, PM_10_, SO_2_, CO, NO_2_, and O_3_ for each city were obtained from air quality stations across China from January 2015 to March 2020. For each city, the average concentration for each pollutant before the COVID-19 outbreak (January 2020) was calculated across the whole period available.

### Statistical methods

To quantify the effect of air pollution on COVID-19 risk, we used the historical data for air quality between 2015 and 2019 and COVID-19 case reports. Socio-demographic and behavioural confounders were identified through the literature search. The association between long-term exposure to air pollutants and COVID-19 risk was assessed by regression with a generalized linear model (GLM):

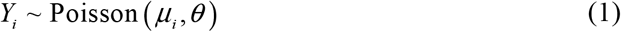

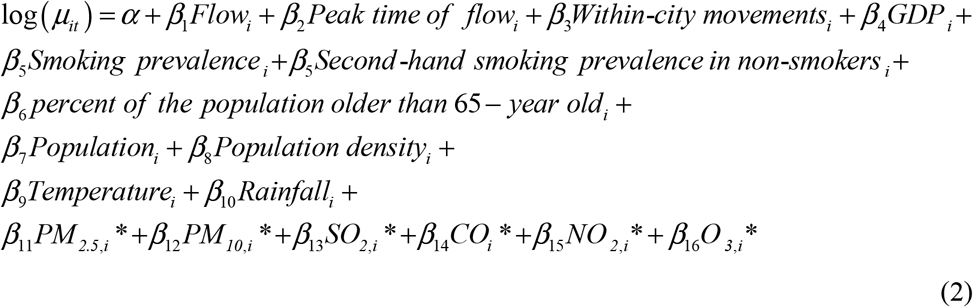

where *Flow_i_* is the passenger volume from Wuhan to city *i* during the Spring Festival 2020, before the Wuhan travel ban, and *Peak time of flow* reflects the corresponding peak time. *Within-city movementsi* shows the effect of social distancing, such as suspending intra-city public transport, closing entertainment venues and banning public gatherings, and was measured by daily human mobility within a city between the travel ban and 6 March 2020 of city *i*. These data were obtained from the Baidu location-based service. GDP is the gross domestic product per capita of city *i. Smoking prevalence* and *second-hand smoking prevalence in non-smokers* were obtained from the Chinese National Nutrition and Health Survey (NNHS) ^14,15^. The proportion of residents older than 65, population and population density of city *i* were extracted from census data. Climate conditions were shown by *temperature* and *rainfall* in summer and winter. *Latitude* and *longitude* show the spatial distribution of city *i*. PM_2.5_, *PM*_10_, *SO*_2_, *CO, NO*_2_, and *O*_3_ show daily average concentration data of air pollutants of city *i* between 1 January 2015 and 31 December 2019. Air pollution variables were included in the model separately because of the high multicollinearity among them. *β*s are regression coefficients. The analysis used the R software (R Foundation for Statistical Computing, version 3.6.3), MASS package.

### Sensitivity analysis

We conducted additional sensitivity analyses to assess the robustness of our results. First, we fit models omitting adjusted variables, separately. We then treated NO_2_ and PM_2.5_ as a categorical variable (categorized at empirical quintiles), then used air pollutant concentrations in winter and non-winter to replace the annual average. Finally, cities were categorized by population size into three separate groups, small (0–2.68 million population, n = 98), medium-sized (2.68–4.67 million, n = 97), and large (4.67–30.75 million, n = 98).

## Results

### Analysis of COVID-19 in China

We used a well-tested research data platform that gathers nationwide human mobility, census, air pollution and socio-economic data, and data on smoking from a nationwide survey ^16^. The records we used consisted of reported COVID-19 cases for 324 cities (except Wuhan city) in mainland China between 31 December 2019 and 6 March 2020 (excluding newly-reported locally-acquired infections), mobile data of recorded movements (data provided by Baidu location-based services and telecommunications operators in China), census data (population, population density, and percentage of the population over 65 years old), smoking prevalence and second-hand smoking prevalence in non-smokers, gross domestic product (GDP) per capita, climate condition (temperature and rainfall), and six major air pollutant concentrations (PM_2.5_, PM_10_, SO_2_, CO, NO_2_, and O_3_). We analysed and quantified the relationship between long-term exposure to air pollution and COVID-19 risk.

Between 31 December 2019 and 6 March 2020, 81,132 cases of COVID-19 were reported across China. Of these, 62.6% (50783/81132) of cases were clustered in Wuhan city, and the remaining 37.4% (30349/81132) cases were distributed across 324 other cities. After 6 March 2020, there were very few locally-acquired infections outside Wuhan city in the first wave. There was sustained local transmission of COVID-19 in Wuhan city, so data from there were not included in the subsequent analysis.

COPD, CVD and hypertension are associated with high risk of COVID-19 severity. We therefore suspected that long-term exposure to air pollutants, which increases lung damage, would be associated with increased vulnerability to COVID-19. To test this hypothesis, socio-demographic and behavioural confounders were identified through a literature search. We first investigated the effect of travel movements from Wuhan city, the location where COVID-19 was first recorded, and from where it spread across China. In 2020, during travel for the Spring Festival holiday, approximately 4.3 million people travelled out of Wuhan to other cities in China ^16^ This travel is strongly associated with the total number of cases reported from each city (rho = 0.81, *P* < 0.01), suggesting the outbreaks across China were mainly seeded from Wuhan city.

### Air pollution reduction and travel restrictions

On 23 January 2020, in an attempt to contain the epidemic, non-essential travel was prohibited in and out of Wuhan city, a major transport hub and conurbation of 11 million people. Since then, the whole of China has implemented the highest level of emergency response to reinforce the containment of COVID-19. Interventions included the closure of entertainment venues, the suspension of within-city public transport, and prohibition of travel to and from other cities across China. Our results indicate that these measures have significantly reduced the movement within cities compared with 2019 (Figure 2). The average air quality also significantly improved, compared with the same period in 2019. The changes in average daily PM_2.5_, PM_10_, SO_2_, CO, NO_2_, and O_3_ were −7.02%, −19.25%, −15.06%, −5.46%, −20.17% and 5.01% from 31 December to 6 March (Figure 2).

**Figure 2.**
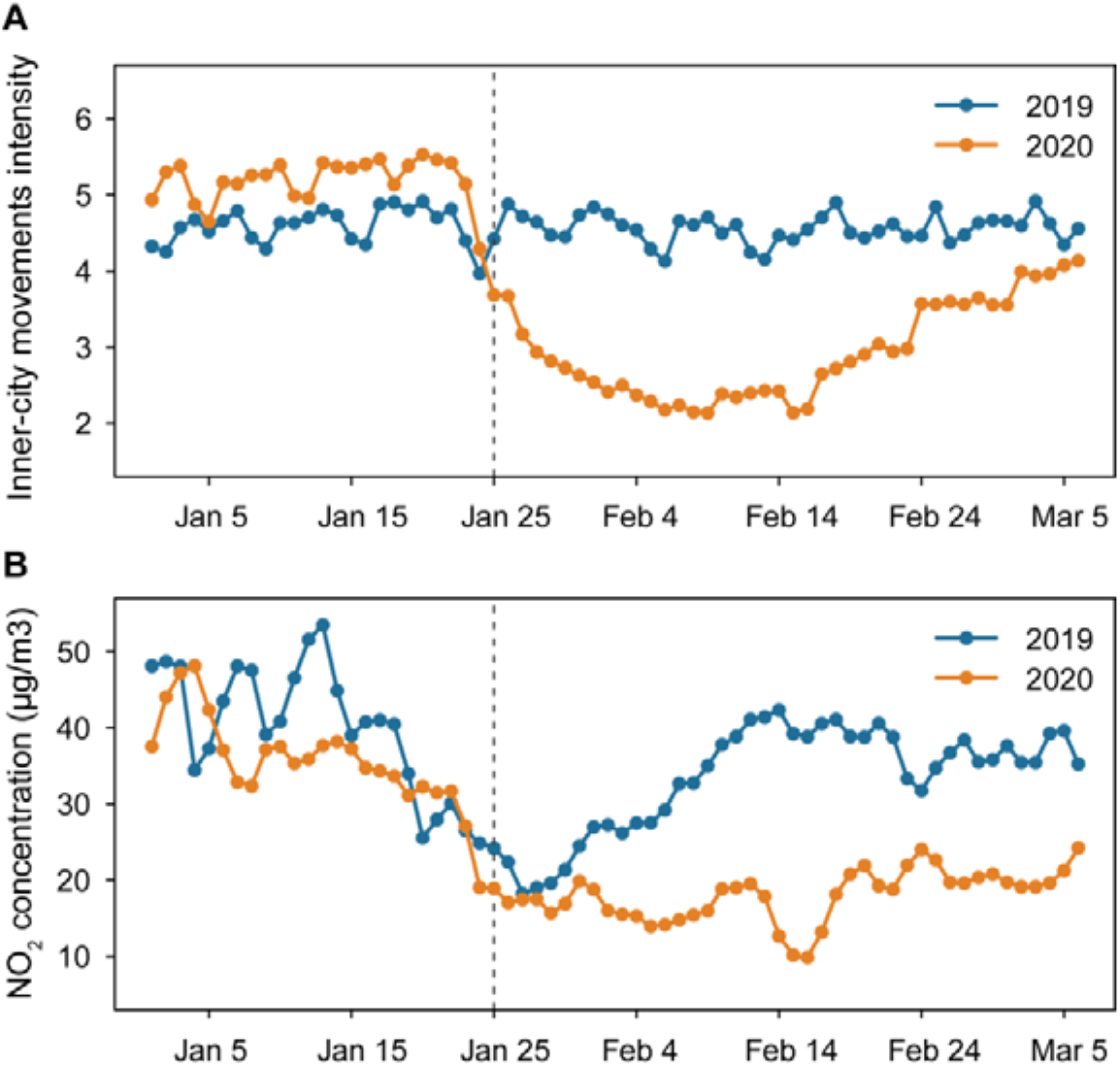
(A) Average within-city movement intensity and (B) air pollutant concentration in 324 cities in China during the 2020 COVID-19 outbreak (orange line), compared with the same period in 2019 (blue line).

### The effect of long-term exposure to NO_2_ and PM_2.5_ on COVID-19 cases and severe infections

We also collected data on a range of confounding variables such as gross domestic product per capita, smoking prevalence, climate, and age composition (age > 65), together with travel movements from Wuhan and within-city movements (as a measure of social distancing). These were used as control variables in the statistical analysis. The population size of each city was not included in the model because of high multicollinearity with travel movements from Wuhan (rho = 0.71, *P* < 0.01). The three billion Spring Festival holiday trips also induced uncoordinated changes in actual population size across cities ^17^ As expected, the number of COVID-19 cases in each city increased with passenger flow from Wuhan. More infections were reported in cities that had more travellers from Wuhan. Overall, we observed positive and significant associations between both reported and severe cases of COVID-19 with historical air pollutant concentration (Figure 3). In 324 cities (except Wuhan) that had data on air quality, an increase of 10 *μ*g/m^3^ in NO_2_ or PM_2.5_ concentration was associated with 22.41% (95% CI: 7.28%–39.89%) or 15.35% (95% CI: 5.60%–25.98%) increase in COVID-19 cases. We also examined the relationship between the number of severe COVID-19 cases and air pollutants. An increase of 10 *μ*g/m^3^ in NO_2_ or PM_2.5_ concentration was associated with a 19.20% (95% CI: 4.03%–36.59%) or 9.61% (95% CI: 0.12%-20.01%) increase in severe COVID-19 cases. The results are statistically significant and robust to sensitivity analyses (Tables 1–3). The analysis was also carried out for separate datasets. Cities were categorized by population size into small, medium, and large. There was a positive but not significant association between the number of confirmed cases and NO_2_ or PM_2.5_ concentration in each group (Figure 3). The results might still be affected by population size, however, because air pollution is usually the result of population growth, so there is a predefined correlation between air pollutant concentration and population size.

**Table 1.** Impact of historic air pollution exposure on COVID-19 risk.

| Covariates | Confirmed COVID-19 cases |  |  | Severe COVID-19 cases |  |  |
| --- | --- | --- | --- | --- | --- | --- |
|  | Coefficient | std | P | Coefficient | std | P |
| **Intercept** | 2.630 | 0.538 | <0.001 | 0.769 | 0.509 | 0.132 |
| **Within-city movements** | -1.065 | 0.139 | <0.001 | -0.778 | 0.137 | <0.001 |
| **Percent of population older than 65** | 0.069 | 0.032 | 0.030 | -0.054 | 0.035 | 0.129 |
| **Inflow from Wuhan** | 0.012 | 0.001 | <0.001 | 0.030 | 0.009 | <0.01 |
| **Peak of inflow from Wuhan** | 0.108 | 0.020 | <0.001 | 0.074 | 0.023 | <0.01 |
| **Mean Temperature of Coldest Quarter** | 0.026 | 0.009 | <0.01 | 0.027 | 0.007 | <0.001 |
| **NO<sub>2</sub>\*** | 0.020 | 0.007 | <0.01 | 0.018 | 0.007 | <0.01 |
| **PM<sub>2.5</sub>\*** | 0.014 | 0.004 | <0.01 | 0.009 | 0.004 | 0.034 |
| **PM<sub>10</sub>\*** | 0.002 | 0.003 | 0.365 | 0.001 | 0.002 | 0.733 |
| **SO<sub>2</sub>\*** | -0.016 | 0.008 | 0.050 | -0.006 | 0.006 | 0.344 |
| **CO\*** | -0.013 | 0.246 | 0.958 | -0.049 | 0.212 | 0.817 |
| **O<sub>3</sub>\*** | -0.006 | 0.005 | 0.249 | -0.010 | 0.005 | 0.049 |

\*variables were included in the model separately because of the high multicollinearity among air pollutants.

**Table 2.**
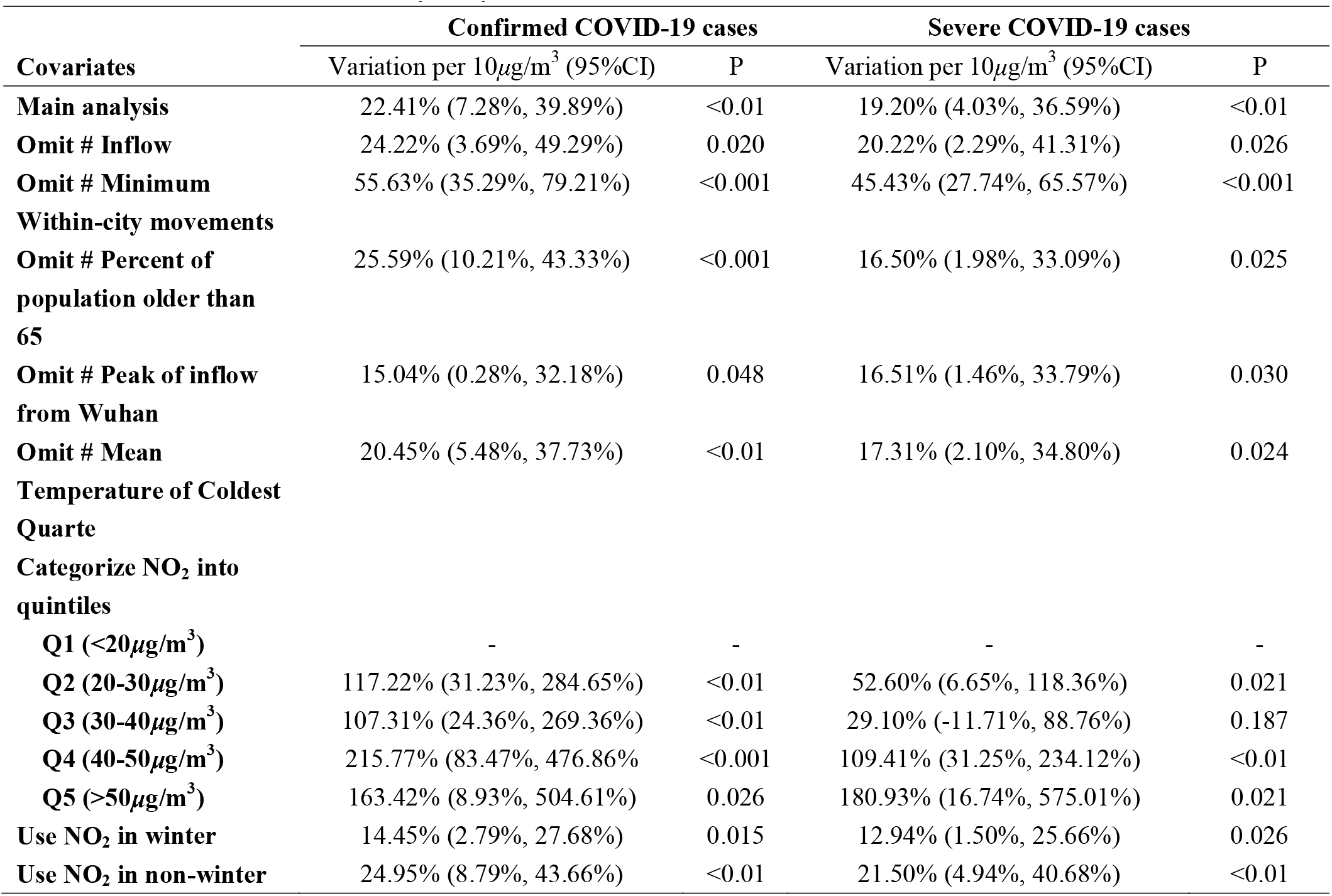
NO_2_-sensitivity analysis.

**Table 3.**
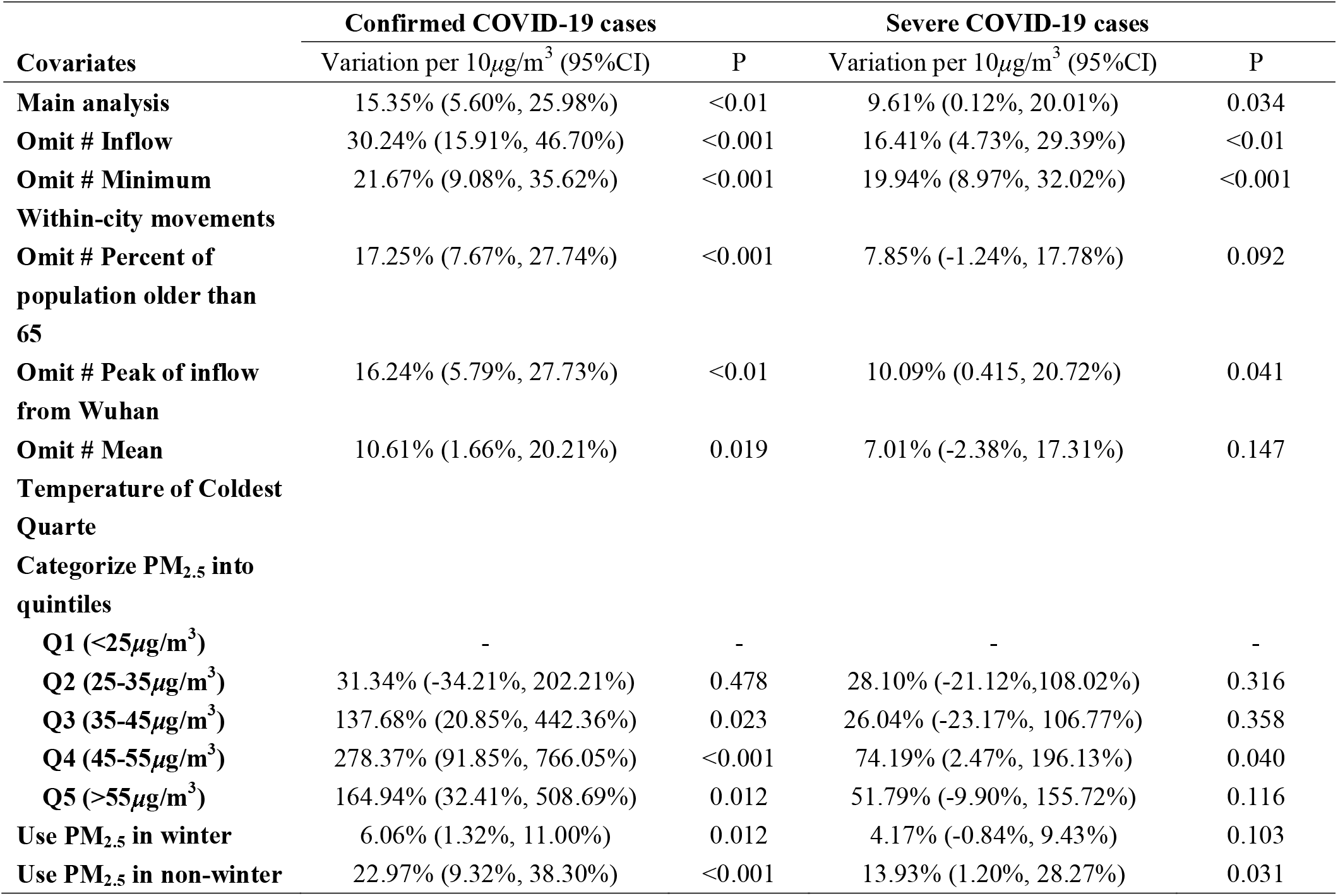
PM_2.5_-sensitivity analysis.

**Figure 3.**
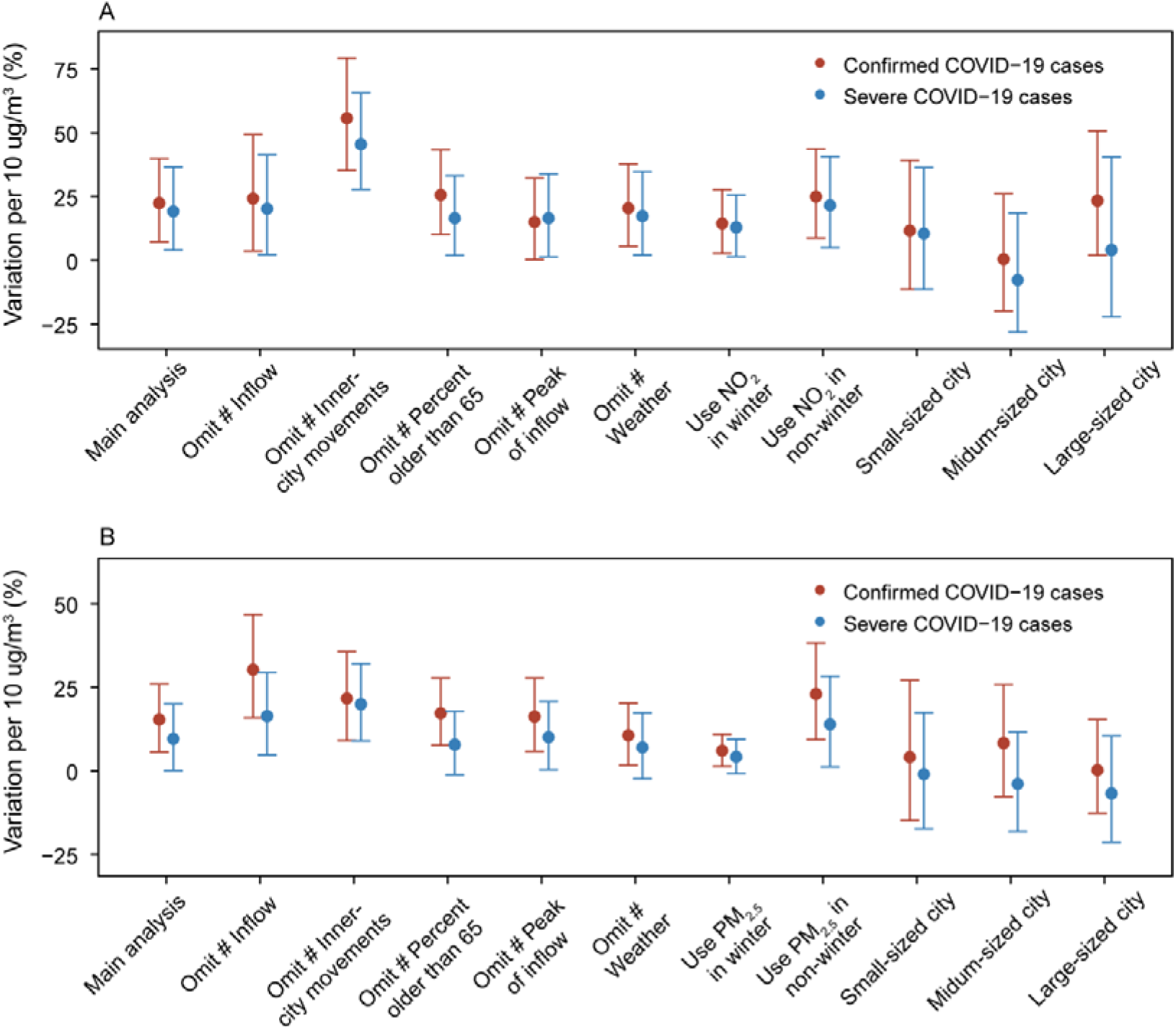
**Variation per unit and 95% confidence intervals, (A) NO_2_ and (B) PM_2.5_**.The variation per unit (VPU) = [exp(variable coefficient) – 1] ×100%. VPU can be interpreted as percentage increase in the number of COVID-19 cases associated with a 10 μg/m^3^ increase in long-term average NO_2_ and PM_2.5_. The VPU from the main analysis was adjusted by confounding factors (inward movement from Wuhan city before travel ban and peak time of movement, within-city movements, GDP, smoking prevalence, second-hand smoking prevalence in non-smokers, percent of the population > 65 years old, population, population density, climate, and spatial distribution). In the sensitivity analyses, we omitted each confounding factor separately, and used seasonal air pollutant concentrations.

## Discussion

Several studies have evaluated the association between air pollution exposure and COVID-19 risk. Our finding is in line with studies based on large-scale nationwide data, including in the US ^18^, UK ^19,20^, Italy ^21,22^, The Netherlands ^23^, Spain, France and Germany ^24^, and provinces of China ^25^. However, the size of the study and that the first wave has been contained in China, meant that we were able to investigate the impact of historical air pollution on COVID-19 risk and severity. Our results also highlighted the importance of air quality improvements to health in China.

Our findings are consistent with previous results that exposure to air pollutants is associated with poorer lung function, often measured by forced vital capacity and forced expiratory volume in 1 second ^26-29^. Particulate matter (PM) is associated with increased risk of cardiopulmonary diseases^30^, aggravated case fatality of severe acute respiratory syndrome (caused by another type of coronavirus) 31 and could impair the immune response^32^. Oxidant pollutants could also damage the immune function and affect the ability of the lungs to clear the virus^6^. Nitrogen oxides (NO_X_) can cause inflammatory responses and have a direct effect on the risk of respiratory diseases^33,34^ (also shown in Comparative Toxicogenomics Database, http://ctdbase.org). Nitrogen oxide is primarily produced by traffic and factories, and less influenced by climate condition, so there were reductions in both the concentration of nitrogen oxides and within-city movements during the COVID-19 outbreak in cities of China. Increased ozone or sulphur dioxide concentrations were associated with lower COVID-19 risk, which is also consistent with previous studies ^19,25^. However, the mechanisms underlying the impact on COVID-19 risk remain unknown.

Several important caveats are worth mentioning. First, the data included here were all from mainland China. It is therefore not clear whether the findings can be generalized to other countries without data on historic air pollution exposure. Second, there are currently no high-quality records at city level of severe COVID-19 infections and ICU admissions, although we have attempted to fill the gap by using province-level reports. Finally, we do not know the exact number of cases because of the number of asymptomatic and mild-symptomatic cases that may not be recorded. These data will not be available until there has been a systematic survey of infection (e.g. by serological testing) across China. However, we have reported a national-level disease pattern covering 324 cities and its potential association with long-term exposure to air pollutants.

In conclusion, our results, combined with recent reports from elsewhere, suggest a potential association between air quality and population vulnerability to COVID-19. Interventions to contain the COVID-19 outbreak in China successfully reduced air pollution levels and potentially prevented acute respiratory disease. The findings of this paper (and other previous studies that have given ambiguous results) indicate that a more definitive analysis is needed of the link between COVID-19 and air pollution.

## Data Availability

We used a well-tested research data platform that gathers nationwide human mobility, census data, air pollution data, smoking data from a nationwide survey, and socio-economic data.

## Acknowledgements

We thank the thousands of CDC staff and local health workers in China who collected data and continue to work to contain COVID-19 in China and elsewhere. Funding for this study was provided by the Research Council of Norway contributed to this work through the COVID-19 Seasonality Project (312740); Beijing Science and Technology Planning Project (Z201100005420010); National Natural Science Foundation of China (91643203, 81673234); Beijing Natural Science Foundation (JQ18025); Beijing Advanced Innovation Program for Land Surface Science; Young Elite Scientist Sponsorship Program by CAST (YESS) (2018QNRC001); H.T., M.U.G.K., and C.D. acknowledge support from the Oxford Martin School; HT acknowledges support from the Military Logistics Research Program. The funders had no role in study design, data collection and analysis, the decision to publish, or in the preparation of the manuscript.

## Author contributions

P.Z., H.T., G.J., N.C.S., and C.D. designed the study. H.S. collected and processed the LBS data. Y. L., X.Y., and B. L. collected the statistical data. Y.L. and C.-H.W. conducted the analyses. H.T., C.-H.W., M.U.G.K., Y.Z., and C.D. edited the manuscript. P.Z. wrote the manuscript. All authors read and approved the manuscript.

